# Genomic characterization of respiratory syncytial virus 2022-2023 outbreak in Washington State, USA

**DOI:** 10.1101/2022.12.12.22283375

**Authors:** Stephanie Goya, Jaydee Sereewit, Daniel Pfalmer, Tien V. Nguyen, Shah A. K. Mohamed Bakhash, Elizabeth B. Sobolik, Alexander L. Greninger

**Affiliations:** Department of Laboratory Medicine and Pathology, University of Washington, Seattle, WA, USA; Vaccine and Infectious Disease Division, Fred Hutchinson Cancer Research Center, Seattle, WA, USA

**Keywords:** respiratory syncytial virus, human orthopneumovirus, genome, COVID-19, evolution, genotype, Washington, USA

## Abstract

Mitigation measures against the COVID-19 pandemic affected the RSV seasonality and led to an unexpectedly high number of RSV cases in Washington State since October 2022. Here we describe the RSV genomic characteristics and evolutionary relationship of 2022 outbreak compared to the previous RSV outbreaks in the region and globally.

## Research Letter

Respiratory syncytial virus (RSV) annual seasonality in Washington State (WA), USA has usually been limited to late-autumn and winter (1). However, no RSV outbreak was detected during the 2020-21 season due to the COVID-19 pandemic. After the lockdown relaxation in summer 2021, an early RSV season started in August (Figure A). The 2022-2023 outbreak also started earlier but unexpectedly, the number of RSV cases was significantly higher than 2021, alarming public health authorities and the general community (2).

**Figure 1.**
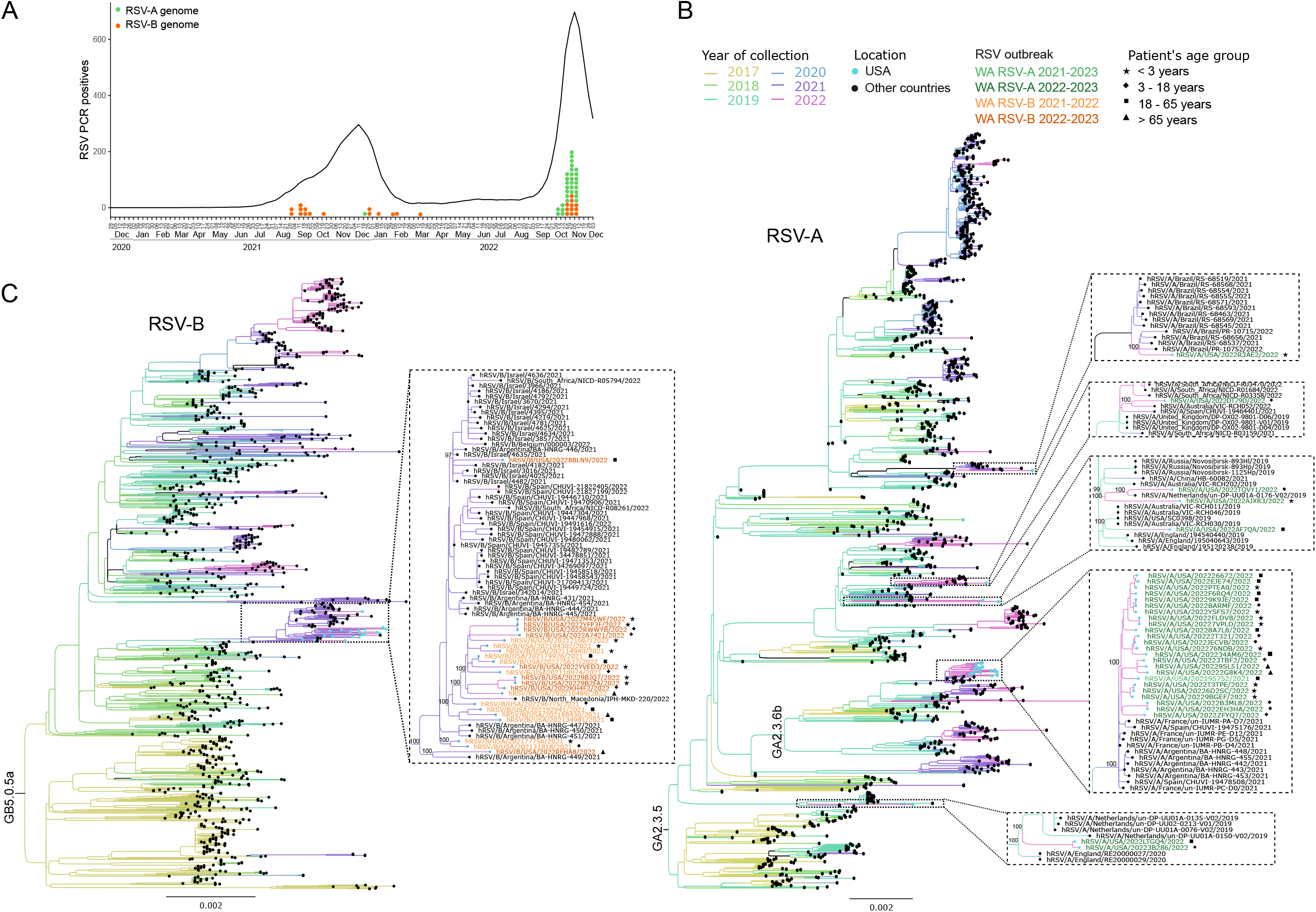
Phylogenetic analysis of RSV in Washington State 2021-2022. A) Weekly WA PCR RSV detections by 5-week average is depicted using data from National Respiratory and Enteric Virus Surveillance System from the National Center for Immunization and Respiratory Diseases, Division of Viral Diseases, reviewed up to 07 Dec 2022. Orange and green dots show the collection date RSV genomes analyzed in this study. Maximum likelihood trees for RSV-A (B) and RSV-B (C) complete genomes collected from 2017 to 2022. Details on the phylogenetic association of the WA genomes are shown in larger size including the UF-bootstrap for the main phylogenetic clades. Lineages GA2.3.5 and GA2.3.6b in RSV-B and GB5.0.5a in RSV-B are labeled in their ancestral nodes. The collection year of the specimens is depicted by tree branch color. RSV genomes from the USA are highlighted with a light blue circle at the tip. WA RSV genomes from the epidemic seasons 2021-2022 and 2022-2023 are highlighted in shades of green for RSV-A and orange for RSV-B. In the larger size panels the age group for each patient is detailed with a symbol detailed in the legend. The scale bar represents the inferred number of nucleotide substitutions per site.

This severe RSV outbreak may be due to diminished protective immunity in the population because of a prolonged period of low exposure to the pathogen (3). There is also uncertainty whether selective pressure due to the low transmission in 2020 has caused the emergence of new viral strains with improved fitness. Here, we evaluated whether the RSV causing the current outbreak has any distinctive genomic characteristics compared to strains from prior seasons.

We performed metagenomic next-generation and hybridization capture-based sequencing of 54 RSV genomes, including 14 RSV strains collected from 2021-2022 and 40 from the 2022-2023 outbreaks in Seattle, WA. Briefly, viral RNA was extracted from excess nasal and nasopharyngeal swab specimens collected from individuals presenting to UW Medicine COVID-19 collection sites, clinics, emergency rooms, and inpatient facilities that tested positive for RSV with a Ct value <30 (Table) (4). All individuals were outpatients except for two hospitalized patients from 2021. For phylogenetic analyses, complete genomes from both subtypes, RSV-A and RSV-B, were downloaded from NCBI GenBank and GISAID public databases. Alignments were built with MAFFT, and phylogenetic trees were inferred with IQ-TREE (5). Additional methodological details are available in Appendix 1. This study was approved by the UW Institutional Review Board with a consent waiver (STUDY00000408).

Among the sequenced specimens, 1 RSV-A and 13 RSV-B were detected during the 2021-2022 season, while 30 RSV-A and 10 RSV-B were detected during the 2022-2023 season (Table 1). No difference in the subtype predominance either by patient’s age group or gender was detected in the 2022-2023 outbreak (Fisher’s exact test, p> 0.1). Genotyping showed that 7 RSV-A sequences classified as GA2.3.5 and 24 as GA2.3.6b genotypes (both comprising ON1 strains) and all RSV-B classified in the GB5.0.5.a genotype (BA strains) (6) (Appendix). Phylogenetic analysis with all RSV genomes available in public databases up to December 2022 showed that WA RSV genomes are closely related with contemporary viruses (Appendix). Reduced phylogenetic trees including all RSV genomes collected from 2017 to 2022 are shown in Figure B/C. In both RSV-A and RSV-B trees, WA genomes from 2021-2022 and 2022-2023 outbreaks were closely related. However, these sequences were not phylogenetically close with previous WA RSV genomes from 2018 and 2019. Some WA 2022 sequences were individually associated with viruses from France, Spain, Argentina, Brazil, Netherlands, Israel, Australia and North Macedonia dating from 2019, 2021 or early-2022, suggesting multiple viral introductions to the region. Nevertheless, most of analyzed WA genomes were associated in a statistically supported monophyletic clade (Figure. B and C). This result indicates that the 2022-23 WA outbreak is mainly caused by the same RSV-A and RSV-B lineages seen globally for approximately a decade. No phylogenetic relationship was seen between sequence clade and patient age.

**Table 1.** Descriptive epidemiology and subtypes of RSV specimens sequenced from the 2021-2022 and 2022-2023 outbreaks in Washington State.

|  | 2021-2022 |  |  | 2022-2023 |  |  |
| --- | --- | --- | --- | --- | --- | --- |
|  | RSV-A | RSV-B | Total | RSV-A | RSV-B | Total |
| Complete genomes | 1 | 13 | 14 | 30 | 10 | 40 |
| Male |  | 6 | 6 | 13 | 7 | 20 |
| Female | 1 | 7 | 8 | 16 | 4 | 20 |
| Clinical status |  |  |  |  |  |  |
| Inpatient | 1 | 1 | 2 |  |  |  |
| Outpatient |  | 12 | 12 | 30 | 10 | 40 |
| Age group |  |  |  |  |  |  |
| <3 years |  | 5 | 5 | 10 | 6 | 16 |
| 3 – 18 years |  | 3 | 3 | 10 | 2 | 12 |
| 19 – 65 years | 1 | 4 | 5 | 8 | 1 | 9 |
| >65 years |  | 2 | 2 | 2 | 1 | 3 |

Analysis of the sequenced 2022-23 WA RSV-A and RSV-B strains by gene showed there had no specific non-synonymous change compared to other RSV strains reported globally. Furthermore, amino acid changes in WA RSV strains were also identified in some sequences from before the pandemic, e.g., the amino acid constellation of A103T and T122A in the RSV-A Fusion protein was also detected in other 14 genomes worldwide, including one sequence from 2019. These results may be a consequence of a bottleneck effect due to the low transmission during 2020 reducing viral diversity (7). An alternating prevalence of RSV subtypes between the previous and current RSV outbreak may also be playing a role in the high levels of viral spread (Table 1). Further studies with more sequences from the region and globally are needed to confirm these hypotheses.

This study is chiefly limited by the low number of RSV genomes from WA during pre-pandemic years and convenience sampling from excess clinical specimens with limited clinical metadata. Nonetheless, WA is comparably a better sampled state for RSV sequences, as only two RSV genomes from the USA since 2017 have been derived from outside of WA.

Several studies have shown the impact on the respiratory virus ecosystem due to the COVID-19 pandemic lockdown measures (8–10). Real-time genomic surveillance of a significant RSV outbreak found no new specific viral changes since the start of the pandemic that account for the increased viral spread. Our data informs the viral evolution of the reemergence of RSV in WA that will help to understand the complex relationship between herd immunity and viral evolution in the post-pandemic era.

### Disclaimers

ALG reports contract testing from Abbott, Cepheid, Novavax, Pfizer, Janssen and Hologic and research support from Gilead and Merck, outside of the described work.

## Data Availability

RSV consensus genomes are available at NCBI GenBank under the accession numbers OP890312 - OP890350 and OP965698 - OP965712. Sequencing reads are available associated with NCBI BioProject PRJNA907066.

https://github.com/greninger-lab/RSV-WA-2022

