## Appendix for "Genomic characterization of respiratory syncytial virus 2022-2023 outbreak in Washington State, USA"

**Material and methods**

*RSV genome sequencing*

Viral RNA was extracted using Quick-RNA Viral Kits (Cat# R1035, Zymo Research). For samples with RSV Ct values below 25, viral genome sequencing was performed as previously described (1). Briefly, extracted RNA was treated with TURBO DNA-free Kit (Cat # AM1907, Thermo Fisher) to remove genomic DNA. First strand cDNA synthesis was performed using random hexamers and SuperScript IV Reverse Transcriptase (Cat# 18090200, Invitrogen); after which, second strand synthesis was performed using Sequenase Version 2.0 DNA Polymerase (Cat# 70775Z1000UN, Applied Biosystems). The double-stranded cDNA was then cleaned up using AMPure XP Magnetic Beads (Cat# A63881, Beckman Coulter) before proceeding to tagmentation and library preparation using Illumina DNA Prep, (S) Tagmentation Kit (Cat# 20025520, Illumina). For samples with RSV Ct values between 25 to 30 the Illumina RNA Prep with Enrichment, (L) Tagmentation (Cat# 20040536) was used with Respiratory Virus Oligos Panel v2 (Cat# 20044311) for the library generation. DNA libraries were sequenced as 2x100bp and 2X250bp run in a NextSeq500 sequencer.

*Bioinformatic analysis*

Quality of FastQ files was analyzed with FastQC (2). RSV genome assembly was generated using Revica pipeline (https://github.com/greninger-lab/revica). Briefly, adapter trimming and quality filtering was performed with Trimmomatic v0.39 (3). Mapping against a viral genome reference database was performed, followed by one round of mapping against the viral reference with the highest median coverage (RSV-A reference selected: MZ516076.1, RSV-B reference selected: OK649754.1) and two iterations of mapping against the consensus reconstruction. Consensus genome was generated considering a minimum base quality of 20, a minimum depth of coverage of 5X and 60% allele frequency.

*Phylogenetic analysis*

RSV genome alignments were built according to the RSV subtype with MAFFT and visualized with Aliview (4,5). For RSV genotyping, the WA sequences were trimmed to the ectodomain of the G gene and analyzed with ReSVidex online and corroborated by maximum likelihood inference including the RSVA- and RSV-B reference alignments available online (6,7). Genotyping trees are available at https://github.com/greninger-lab/RSV-WA-2022. For comprehensive phylogenetic analyses, all RSV genomes from clinical samples with less than 5% Ns publicly available in GenBank and GISAID databases were downloaded. Maximum likelihood trees were inferred using IQ-TREE v2.1(8). The molecular evolution model was estimated with ModelFinder and the reliability of sequences clusters was evaluated using UFBoot2 method (10000 replicates) (9,10). Complete RSV-A and RSV-B tree files as well as the extended version of the reduced trees showed in Figure 1 are available at https://github.com/greninger-lab/RSV-WA-2022.

*Data availability*

RSV consensus genomes are available at NCBI GenBank under the accession numbers OP890312 - OP890350 and OP965698 - OP965712. Sequencing reads are available associated with NCBI BioProject PRJNA907066. Line-item specimen data is available in the Appendix Table.

**Appendix Table -** Sequenced RSV specimen metadata

| Sequence name | Collection date | RSV subtype | Ct value | Genbank Accession Number | BioProject | BioSample | SRA fastq file |
|---|---|---|---|---|---|---|---|
| hRSV/A/USA/202276NDB/2022 | 10-2022 | A | 19.20 | OP890312 | PRJNA907066 | SAMN32118079 | SRR22580785 |
| hRSV/A/USA/2022FLDV8/2022 | 10-2022 | A | 18.46 | OP890313 | PRJNA907066 | SAMN32118080 | SRR22580784 |
| hRSV/A/USA/2022R3AE2/2022 | 10-2022 | A | 18.65 | OP890314 | PRJNA907066 | SAMN32118081 | SRR22580773 |
| hRSV/A/USA/2022AF7QA/2022 | 10-2022 | A | 19.08 | OP890315 | PRJNA907066 | SAMN32118082 | SRR22580762 |
| hRSV/A/USA/2022LTGQ4/2022 | 10-2022 | A | 19.27 | OP890316 | PRJNA907066 | SAMN32118083 | SRR22580751 |
| hRSV/A/USA/202226672/2022 | 10-2022 | A | 20.46 | OP890317 | PRJNA907066 | SAMN32118084 | SRR22580740 |
| hRSV/B/USA/20229B2FA/2022 | 11-2022 | B | 19.33 | OP890341 | PRJNA907066 | SAMN32118085 | SRR22580735 |
| hRSV/A/USA/20223TBF2/2022 | 10-2022 | A | 19.19 | OP890318 | PRJNA907066 | SAMN32118086 | SRR22580734 |
| hRSV/A/USA/2022TQVY1/2022 | 11-2022 | A | 18.78 | OP890319 | PRJNA907066 | SAMN32118087 | SRR22580733 |
| hRSV/A/USA/20229K9JE/2022 | 11-2022 | A | 19.78 | OP890320 | PRJNA907066 | SAMN32118088 | SRR22580732 |
| hRSV/A/USA/20222G8K4/2022 | 10-2022 | A | 17.13 | OP890321 | PRJNA907066 | SAMN32118089 | SRR22580783 |
| hRSV/A/USA/2022DT79D/2022 | 10-2022 | A | 18.84 | OP890322 | PRJNA907066 | SAMN32118090 | SRR22580782 |
| hRSV/B/USA/20229BJQ7/2022 | 11-2022 | B | 18.79 | OP890342 | PRJNA907066 | SAMN32118091 | SRR22580781 |
| hRSV/A/USA/202234AM6/2022 | 11-2022 | A | 18.82 | OP890323 | PRJNA907066 | SAMN32118092 | SRR22580780 |
| hRSV/A/USA/2022YSFS7/2022 | 10-2022 | A | 18.47 | OP890324 | PRJNA907066 | SAMN32118093 | SRR22580779 |
| hRSV/B/USA/2022A7421/2022 | 10-2022 | B | 19.68 | OP890343 | PRJNA907066 | SAMN32118094 | SRR22580778 |
| hRSV/A/USA/2022PTEA0/2022 | 10-2022 | A | 19.11 | OP890325 | PRJNA907066 | SAMN32118095 | SRR22580777 |
| hRSV/B/USA/2022KH4F2/2022 | 10-2022 | B | 19.98 | OP890344 | PRJNA907066 | SAMN32118096 | SRR22580776 |
| hRSV/B/USA/2022RWWYB/2022 | 10-2022 | B | 18.43 | OP890345 | PRJNA907066 | SAMN32118097 | SRR22580775 |
| hRSV/A/USA/20227VPLD/2022 | 10-2022 | A | 18.83 | OP890326 | PRJNA907066 | SAMN32118098 | SRR22580774 |
| hRSV/A/USA/2022T3TPE/2022 | 10-2022 | A | 19.26 | OP890327 | PRJNA907066 | SAMN32118099 | SRR22580772 |
| hRSV/A/USA/2022BARMF/2022 | 11-2022 | A | 17.97 | OP890328 | PRJNA907066 | SAMN32118100 | SRR22580771 |
| hRSV/A/USA/20222T321/2022 | 11-2022 | A | 17.36 | OP890329 | PRJNA907066 | SAMN32118101 | SRR22580770 |
| hRSV/B/USA/20228BLN9/2022 | 10-2022 | B | 20.3 | OP890346 | PRJNA907066 | SAMN32118102 | SRR22580769 |
| hRSV/B/USA/2022RFHA8/2022 | 11-2022 | B | 19.59 | OP890347 | PRJNA907066 | SAMN32118103 | SRR22580768 |
| hRSV/A/USA/2022ZFYQ7/2022 | 11-2022 | A | 22.53 | OP890330 | PRJNA907066 | SAMN32118104 | SRR22580767 |
| hRSV/A/USA/20226D2SC/2022 | 10-2022 | A | 17.85 | OP890331 | PRJNA907066 | SAMN32118105 | SRR22580766 |
| hRSV/A/USA/20228A7L8/2022 | 10-2022 | A | 19.74 | OP890332 | PRJNA907066 | SAMN32118106 | SRR22580765 |
| hRSV/A/USA/2022B3ML8/2022 | 11-2022 | A | 18.1 | OP965712 | PRJNA907066 | SAMN32118107 | SRR22580764 |
| hRSV/A/USA/2022EH3HA/2022 | 11-2022 | A | 20.2 | OP890333 | PRJNA907066 | SAMN32118108 | SRR22580763 |
| hRSV/A/USA/20229BGEF/2022 | 11-2022 | A | 20.26 | OP890334 | PRJNA907066 | SAMN32118109 | SRR22580761 |
| hRSV/A/USA/2022EJE74/2022 | 11-2022 | A | 17.89 | OP890335 | PRJNA907066 | SAMN32118110 | SRR22580760 |
| hRSV/A/USA/2022F6RQ4/2022 | 11-2022 | A | 17.88 | OP890336 | PRJNA907066 | SAMN32118111 | SRR22580759 |
| hRSV/A/USA/20223B286/2022 | 11-2022 | A | 19.46 | OP890337 | PRJNA907066 | SAMN32118112 | SRR22580758 |
| hRSV/B/USA/2022YVED3/2022 | 11-2022 | B | 19.46 | OP890348 | PRJNA907066 | SAMN32118113 | SRR22580757 |
| hRSV/A/USA/20229SL51/2022 | 10-2022 | A | 19.80 | OP890338 | PRJNA907066 | SAMN32118114 | SRR22580756 |
| hRSV/A/USA/2022JECVB/2022 | 10-2022 | A | 19.58 | OP890339 | PRJNA907066 | SAMN32118115 | SRR22580755 |

| | | | | | | | |
|---|---|---|---|---|---|---|---|
| hRSV/B/USA/2022YFP3F/2022 | 10-2022 | B | 19.09 | OP890349 | PRJNA907066 | SAMN32118116 | SRR22580754 |
| hRSV/B/USA/2022M4SWF/2022 | 10-2022 | B | 18.65 | OP890350 | PRJNA907066 | SAMN32118117 | SRR22580753 |
| hRSV/A/USA/2022AJXR3/2022 | 10-2022 | A | 18.58 | OP890340 | PRJNA907066 | SAMN32118118 | SRR22580752 |
| hRSV/B/USA/202177756/2021 | 08-2021 | B | 19.4 | OP965698 | PRJNA907066 | SAMN32118119 | SRR22580750 |
| hRSV/B/USA/202196775/2021 | 09-2021 | B | 22.9 | OP965699 | PRJNA907066 | SAMN32118120 | SRR22580749 |
| hRSV/B/USA/202118974/2021 | 09-2021 | B | 21.1 | OP965700 | PRJNA907066 | SAMN32118121 | SRR22580748 |
| hRSV/B/USA/202131818/2021 | 09-2021 | B | 22.5 | OP965701 | PRJNA907066 | SAMN32118122 | SRR22580747 |
| hRSV/B/USA/202134981/2021 | 09-2021 | B | 23.9 | OP965702 | PRJNA907066 | SAMN32118123 | SRR22580746 |
| hRSV/B/USA/202179926/2021 | 10-2021 | B | 20.4 | OP965703 | PRJNA907066 | SAMN32118124 | SRR22580745 |
| hRSV/A/USA/202195752/2021 | 12-2021 | A | 18.6 | OP965711 | PRJNA907066 | SAMN32118125 | SRR22580744 |
| hRSV/B/USA/202221067/2022 | 03-2022 | B | 21.6 | OP965704 | PRJNA907066 | SAMN32118126 | SRR22580743 |
| hRSV/B/USA/202210489/2022 | 02-2022 | B | 28.3 | OP965708 | PRJNA907066 | SAMN32118130 | SRR22580738 |
| hRSV/B/USA/202275637/2022 | 02-2022 | B | 26.5 | OP965709 | PRJNA907066 | SAMN32118131 | SRR22580737 |
| hRSV/B/USA/202188430/2021 | 12-2021 | B | 30 | OP965710 | PRJNA907066 | SAMN32118132 | SRR22580736 |

*GISAID acknowledgement*

We gratefully acknowledge the authors from the originating laboratories responsible for obtaining the specimens and the submitting laboratories where genetic sequence data were generated and shared via the GISAID Initiative (Authors acknowledgment table available in https://github.com/greninger-lab/RSV-WA-2022).
